# Genome-wide Analysis Reflects Novel 5-Hydroxymethylcytosines Implicated in Diabetic Nephropathy

**DOI:** 10.1101/2022.01.14.22269066

**Authors:** Ying Yang, Chang Zeng, Kun Yang, Zhou Zhang, Qinyun Cai, Chuan He, Wei Zhang, Song-Mei Liu

## Abstract

Long-term complications of type 2 diabetes (T2D) are the major causes for T2D-related disability and mortality. Notably, diabetic nephropathy (DN) has become the most frequent cause of end-stage renal disease (ESRD) in most countries. Understanding epigenetic contributors to DN can provide novel insights into this complex disorder and lay the foundation for more effective monitoring tools and preventive interventions, critical for achieving the ultimate goal of improving patient care and reducing healthcare burden. We have used a selective chemical labeling technique (5hmC-Seal) to profile genome-wide distributions of 5-hydroxymethylcytosines (5hmC), a gene activation mark, in patient-derived circulating cell-free DNA (cfDNA). Differentially modified 5hmC genes were identified across T2D patients with DN (n = 12), T2D patients with non-DN vascular complications (non-DN) (n = 29), and T2D patients with no complications (controls) (n = 14). Specifically, differential 5hmC markers between DN and controls revealed relevant pathways such as NOD-like receptor signaling pathway and tyrosine metabolism. A ten-gene panel was shown to provide differential 5hmC patterns between controls and DN, as well as between controls and non-DN patients using a machine learning approach. The 5hmC profiles in cfDNA reflected novel DN-associated epigenetic modifications relevant to the disease pathogenesis of DN. Importantly, these findings in cfDNA, a convenient liquid biopsy, have the potential to be exploited as a clinically useful tool for predicting DN in high risk T2D patients.

## INTRODUCTION

Diabetic nephropathy (DN) is one of the most common complications of type 2 diabetes (T2D) and a leading cause for end-stage renal disease (ESRD) globally.(1) Approximately 20% to 40% of T2D patients will ultimately develop nephropathic diseases, thus posing a significant risk for T2D patients.(2) Early detection and preventive intervention of DN has been limited due largely to the lack of comprehensive understanding of its complex pathogenesis and effective biomarkers. Notably, conventional clinical markers to evaluate renal functions of DN, including serum creatinine, estimated glomerular filtration rate (eGFR), and urinary albumin, can be influenced by many factors.(3) Pathologically, the “gold standard” to diagnose DN has been percutaneous renal biopsy. However, various complications can be caused by the procedure, such as bleeding, pain, and infection.(4) Therefore, investigation of novel molecular contributors implicated in DN would not only enhance our understanding of this disease, but also provide opportunities to develop more effective diagnostic and preventive approaches. Of particular interest are novel epigenetic modifications that can be detected and investigated in clinically convenient liquid biopsies, i.e., circulating cell-free DNA (cfDNA), considering their biological relevance in complex disease that reflect systematic changes during pathogenesis.(5)

Particularly, epigenetic modifications are gene regulatory elements that sit between phenotypes and genotypes.(3) To date, the most-investigated epigenetic modification is DNA methylation, i.e., 5-methylcytosines (5mC), which has been implicated in various normal and pathogenesis processes. The regulation of DNA methylation *in vivo* is a dynamic process. The ten-eleven translocation (TET) enzymes can oxidize 5mC into 5-hydroxymethylcytosine (5hmC), 5-aldehyde cytosine (5fC), and 5-carboxylcytosine (5caC) under an active demethylation process.(6) Unlike other demethylated products of 5mC, 5hmC is relatively abundant and biochemically stable in the human genome. Previous studies have confirmed that the 5hmC modification shows a distinct genomic distribution and gene regulatory role from 5mC(7) and has been implicated in a variety of diseases or conditions. Notably, recent studies have begun to demonstrate association of altered 5hmC with diabetes-related conditions such as hyperglycemia.(8)

Technically, the widely-used bisulfite conversion-based epigenomic profiling techniques, though offering opportunities of profiling genome-wide cytosine modifications, cannot distinguish 5hmC from 5mC.(9) Therefore, to investigate the specific distributions of 5hmC in DN, we utilized the 5hmC-Seal technique,(10) a highly sensitive, chemical labeling technique for genome-wide profiling of 5hmC, and the next-generation sequencing (NGS), in cfDNA samples derived from a cohort of T2D patients with and without DN. The 5hmC-Seal technique has been demonstrated by our team and other groups as a reliable approach for biomarker discovery(9, 11-15) using limited clinical biospecimens, e.g., as low as a few nanograms of cfDNA that can be conveniently isolated from 1-2 mL of plasma,(11) a technical advantage especially suitable for future clinical implementation of cfDNA-based non-invasive tools for disease diagnosis and surveillance. Our previous genome-wide analyses of 5hmC suggested a link between altered 5hmC with T2D-assocaited vascular complications in general.(16) For example, the 5hmC-based signatures in cfDNA have been shown to have the potential to distinguish T2D patients with multiple vascular complications from those with single vascular complications,(16) as well as between T2D patients who developed diabetic retinopathy and those who did not.(17) However, whether specific 5hmC changes are implicated in DN has not been investigated yet.

Specifically, in the current study (**Figure 1**), we profiled genome-wide 5hmC in cfDNA samples derived from a cohort of 55 patients with T2D using the 5hmC-Seal technique. Differential analysis was performed to identify DN-associated modified genes as well as involved pathways. We also explored the diagnostic value of 5hmC-based epigenetic signature for DN by investigating differential 5hmC for each patient using a machine-learning approach. Findings from this study suggested an altered 5hmC landscape associated with DN, and provided the foundation for exploiting these novel epigenetic contributors to develop more effective and non-invasive tools for DN diagnosis and preventive intervention in the future.

**Figure 1.**
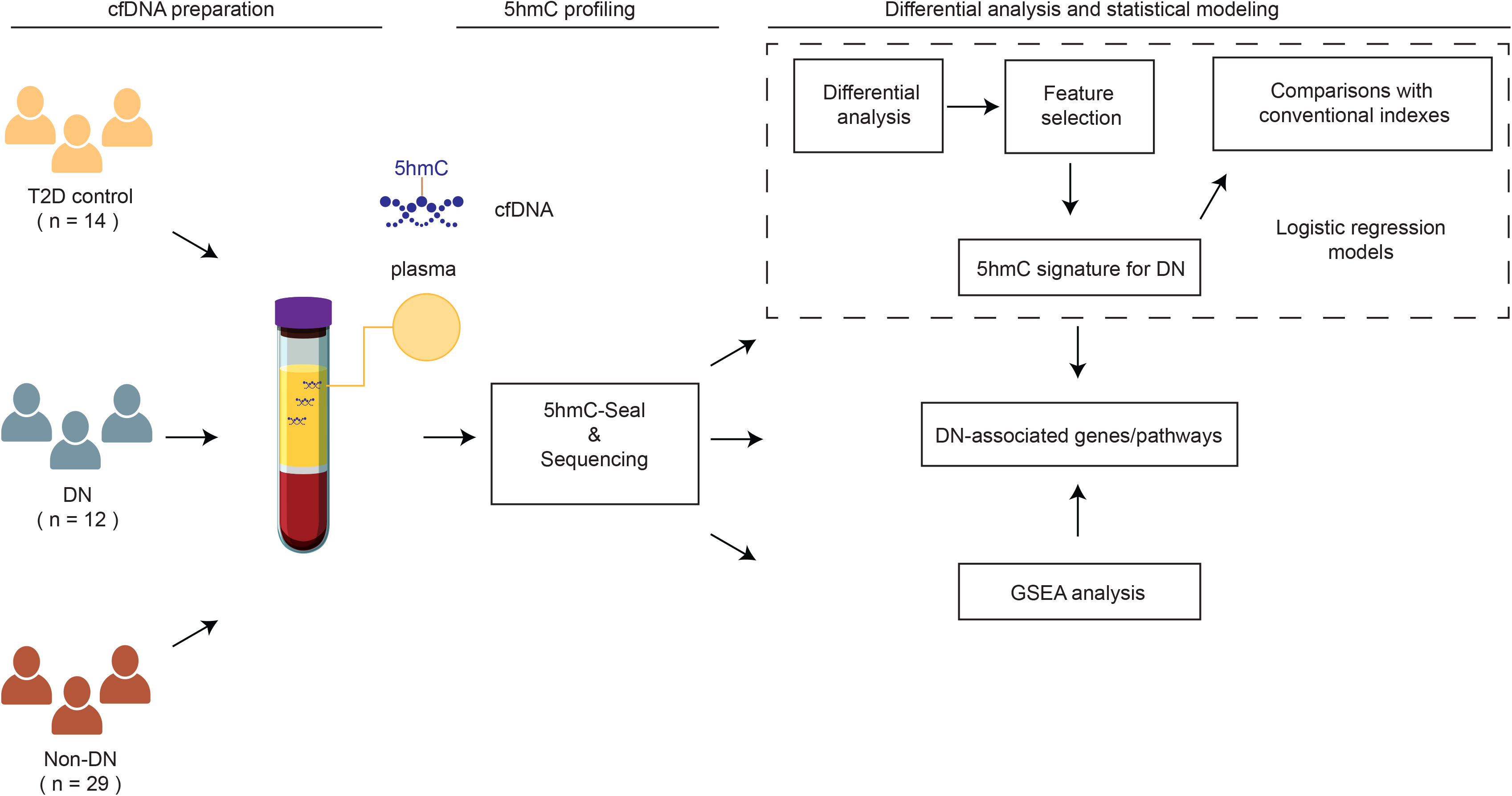
Study design. A cohort of 55 patients with type 2 diabetes (T2D), including 12 patients with diabetic nephropathy (DN), 29 patients with non-DN complications (Non-DN), and 14 controls (CTRL), are profiled for genome-wide 5-hydroxymethylcytosines (5hmC) using the 5hmC-Seal technique and the next-generation sequencing, followed by differential analysis and gene set enrichment analysis (GSEA), feature selection and modeling to inform biological insights and evaluate biomarker potential.

## METHODS

### Study populations

A total of 55 patients with T2D, including 12 patients with DN, 29 patients with non-DN complications (i.e., macrovascular complications, neuropathy, and retinopathy), and 14 gender-, age-matched T2D controls with no complications were recruited at Zhongnan Hospital of Wuhan University, China. Patients were diagnosed according to the 2017 Standards of Medical Care in Diabetes of the American Diabetes Association (ADA).(18) Clinical variables were collected from the medical records following a standard protocol at Zhongnan Hospital. Fasting plasma samples (∼2 mL/patient) were collected at the following morning after hospital admission. This study was approved by the Medical Ethics Committee of Zhongnan Hospital of Wuhan University (2019069). Informed consent was obtained from each study participant.

### Laboratory measurements

Laboratory measurements were obtained when the blood samples were collected at Zhongnan Hospital for the current study. The AU5800 Chemistry Analyzer (Beckman) was used to detect glucose, creatinine (CREA), blood urea nitrogen (BUN), uric acid (UA). The HA-8160 Glycohemoglobin Analyzer (ADAMS) was used to measure blood glycated hemoglobin (HbA1c). Serum insulin was assayed by the i4000SR Immunology Analyzer (Abbott Laboratories). The eGFR was calculated by the Chronic Kidney Disease Epidemiology Collaboration equation.(19)

### Preparation of cfDNA samples, 5hmC-Seal assay, and data processing

The preparation of circulating cfDNA samples, 5hmC-Seal library construction, and data processing has been described in our previous publications.(10, 11) Briefly, plasma samples were separated and stored at –80 °C after centrifuging twice at 1,350× g for 12 min and 13,500× g for 5 min. cfDNA was extracted from the plasma using the Circulating Nucleic Acid Kit (Qiagen, Germany) according to the manufacturer’s instructions. The quality of cfDNA was examined using the standard molecular biology approach. The 5hmC-containing cfDNA fragments were then specifically pulled down and enriched with chemical labeling of 5hmC before being amplified to construct libraries for the next-generation sequencing (NGS). The 5hmC-Seal library construction and the NGS with the Illumina NextSeq 500 platform (paired-end 39 bp) was performed by Innovation Center for Genomics, Peking University (Beijing, China). On average, ∼ 23 million unique reads/sample were obtained from the NGS. The raw 5hmC-Seal data were then processed to accommodate downstream analysis. According to our previous studies, gene bodies (hg19) were our primary targets used to summarize the 5hmC-Seal data from the NGS by counting the sequencing reads. Additionally, kidney-derived histone modification marks for enhancers (H3K4me3, H3K27ac) were obtained from the Roadmap Epigenomics Project(20) to help inform biological insights.

### Identifying DN-associated 5hmC signature in cfDNA

Multivariable logistic regression models were used to identify genes containing differential 5hmC levels (i.e., normalized read counts) between DN patients and T2D controls, as well as between DN and non-DN patients. Though not the focus of the current study, we also performed differential analysis between T2D controls and patients with non-DN complications for comparison. Adjusted covariates included age and gender. Considering the limited sample size in the current study, we used fold change and raw p-value to support statistical significance.

Besides single gene-based differential analysis, the Gene Set Enrichment Analysis (GSEA)(21) was also used to explore functional enrichment of canonical pathways between diagnosis classes, e.g., DN vs. controls using the clusterProfiler tool (v4.0).(22) Specifically, the pathways maintained at the Kyoto Encyclopedia of Genes and Genomes (KEGG)(23) database were evaluated for enrichment in patients with DN relative to either controls or patients with non-DN complications. Genes were ranked by fold change (DN/controls or DN/non-DN) in a decreasing order, and false discovery rate (FDR) was used to identify any enriched pathways (≥ 15 genes and FDR < 0.05) along the ranked gene lists from the GSEA.

### Summarization of a 5hmC-based epigenetic score for DN

To evaluate whether a cfDNA-based score with potential diagnostic value could be summarized from the 5hmC-Seal data, those genes that were differentially modified between 1) DN and controls; 2) DN and non-DN complications; but not in 3) non-DN complications and controls, were further selected to build a signature panel by applying the elastic net regularization(24) on the multivariable logistic regression models. The final signature genes were selected if they were consistently present (100%) in 100 iterations using repeated two-fold cross-validation to differentiate between DN and controls. A weighted score:

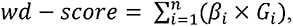

where *G*_*i*_ represents the normalized read count of the *ith* gene body and *β*_*i*_ represents its regression coefficient, was computed, following our previous publications.(11, 12, 16, 17) The area under the receiver operator characteristic (ROC) curve (AUROC) was used to demonstrate model performance. The 95% confidence intervals (CI) were calculated with the DeLong method.(25) The optimal score cutoffs for the AUROCs were determined by the score that maximized the Youden Index, and the corresponding sensitivity and specificity were estimated.

### Comparison between the wd-score for DN with conventional clinical variables or risk factors

To compare the performance of the 5hmC-based scores for DN relative to various clinical variables, univariable logistic regression models for available clinical variables were examined as follows:

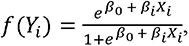

where *Y*_*i*_ represents binary diagnosis classes (i.e., DN vs. non-DN/controls; or DN vs. non-DN), *X*_*i*_ represents age, gender, or each of the following clinical variables, including BMI, smoking history, drinking history, glucose, HbA1c, insulin, CREA, UA, BUN, and eGFR. The predicted probabilities of the univariable logistic regression models were used for assessing classification performance: i.e., DN vs. non-DN/controls; or DN vs. non-DN, via the AUROC. Sensitivity and specificity at the cutoff that maximized the Youden index were estimated for each variable.

## RESULTS

### Clinical and demographic characteristics of the study subjects

**Table 1** shows the clinical and demographic characteristics of the 55 study participants. Overall, there were no significant differences regarding major demographic and clinical variables between patient groups. There were comparable distributions of potential confounders for epigenetic modifications between patient groups, such as baseline BMI (body mass index) and gender (P>0.05). Notably, differences in age at the time of blood collection were observed between T2D controls and DN patients. Therefore, age was used as a covariate in downstream differential analysis when comparing between diagnosis groups (e.g., DN vs. controls). Moreover, totally 24 patients used insulin treatment and 27 patients used oral glucose-lowering medications, though there was no significant disparity regarding medication treatment between different diagnosis groups (P>0.05).

**Table 1.** Demographical and clinical characteristics of the study subjects.

| Clinical Variable | T2D control<br>(n=14) | DN<br>(n=12) | Non-DN<br>(n=29) | P<sup>A</sup> | P<sup>B</sup> | P<sup>C</sup> |
| --- | --- | --- | --- | --- | --- | --- |
| Age (year) | 47.1±9.7 | 56.8±14.5 | 57.7±8.1 | 0.08<sup>a</sup> | 0<sup>a</sup> | 0.71<sup>a</sup> |
| Gender (male/female) | 10/4 | 9/3 | 13/16 | 0.52<sup>b</sup> | 0.14<sup>b</sup> | 0.25<sup>b</sup> |
| BMI (kg/m<sup>2</sup>) | 24.9±3.7 | 25.4±2.3 | 25.7±4.1 | 0.49<sup>a</sup> | 0.62<sup>a</sup> | 0.99<sup>a</sup> |
| Smoking (Yes/No) | 6/8 | 7/5 | 5/24 | 0.69<sup>b</sup> | 0.15<sup>b</sup> | 0.02<sup>b</sup> |
| Drinking (Yes/No) | 2/12 | 4/8 | 4/25 | 0.5<sup>b</sup> | 1<sup>b</sup> | 0.32<sup>b</sup> |
| T2D duration (year) | 3.7±3.9 | 4.5±6.4 | 6.9±6.3 | 0.7<sup>a</sup> | 0.14<sup>a</sup> | 0.08<sup>a</sup> |
| SBP (mmHg) | 123.6±9.4 | 135.4±20.2 | 133.9±19.2 | 0.17<sup>a</sup> | 0.09<sup>a</sup> | 0.82<sup>a</sup> |
| DBP (mmHg) | 78.3±7.9 | 82.1±10.6 | 77.8±10.6 | 0.28<sup>a</sup> | 0.47<sup>a</sup> | 0.13<sup>a</sup> |
| Glucose (mmol/L) | 11.5±3.3 | 9.5±3.5 | 8.5±3.1 | 0.13<sup>a</sup> | 0.01<sup>a</sup> | 0.46<sup>a</sup> |
| HbA1c (%) | 9.1±1.8 | 7.9±2.2 | 8.4±1.9 | 0.17<sup>a</sup> | 0.33<sup>a</sup> | 0.42<sup>a</sup> |
| Insulin (uU/mL) | 10.3±6.0 | 7.4±7.1 | 9.5±7.0 | 0.28<sup>a</sup> | 0.66<sup>a</sup> | 0.28<sup>a</sup> |
| Renal Function: |  |  |  |  |  |  |
| BUN (mmol/L) | 5.8±1.3 | 5.7±1.8 | 5.9±1.7 | 0.78<sup>a</sup> | 0.83<sup>a</sup> | 0.69<sup>a</sup> |
| CREA (umol/L) | 60.3±13.9 | 78.8±31 | 63.1±18.2 | 0.15<sup>a</sup> | 0.86<sup>a</sup> | 0.14<sup>a</sup> |
| UA (umol/L) | 283.6±51.9 | 319.7±99.1 | 291.1±81.9 | 0.4<sup>a</sup> | 0.93<sup>a</sup> | 0.51<sup>a</sup> |
| eGFR (mL/min/1.73m<sup>2</sup>) | 111.6±12.2 | 89.2±28.4 | 98.5±13.4 | 0.02<sup>a</sup> | 0.01<sup>a</sup> | 0.24<sup>a</sup> |
| Medication (Yes/No) |  |  |  |  |  |  |
| Insulin | 6/8 | 3/9 | 15/13 | 0.59<sup>b</sup> | 0.74<sup>b</sup> | 0.19<sup>b</sup> |
| Oral glucose-lowering | 6/8 | 6/6 | 15/13 | 1<sup>b</sup> | 0.74<sup>b</sup> | 1<sup>b</sup> |

Notes: <sup>A</sup>, Comparison between T2D control (CTRL) and Nephropathy (DN); <sup>B</sup>, Comparison between T2D control (CTRL) and patients with non-nephropathy complications (Non-DN); <sup>C</sup>, Comparison between DN and Non-DN; a, Wilcoxon test; b, chi-square test. BMI: Body mass index; SBP: Systolic blood pressure; DBP: Diastolic blood pressure; HbA1c: Glycated hemoglobin; CREA: Creatinine; UA: Uric acid; BUN: Blood urea nitrogen; eGFR: Estimated glomerular filtration rate. T2D: type 2 diabetes; DN: diabetic nephropathy.

### Differentially modified genes associated with DN

Differentially modified genes were detected by pair-wise comparisons using multivariable logistic regression models, controlling age and gender. A total of 336 genes were detected to be differentially modified between T2D controls and patients with DN (P<0.05), among which 271 genes with a fold change of at least 10%. In comparison, 427 genes were found to be differentially modified between patients with DN and patients with non-DN complications (P<0.05), among which 250 genes with a fold change of at least 10%, indicating the presence of 5hmC signatures specific to DN complications (**Figure 2A**).

**Figure 2.**
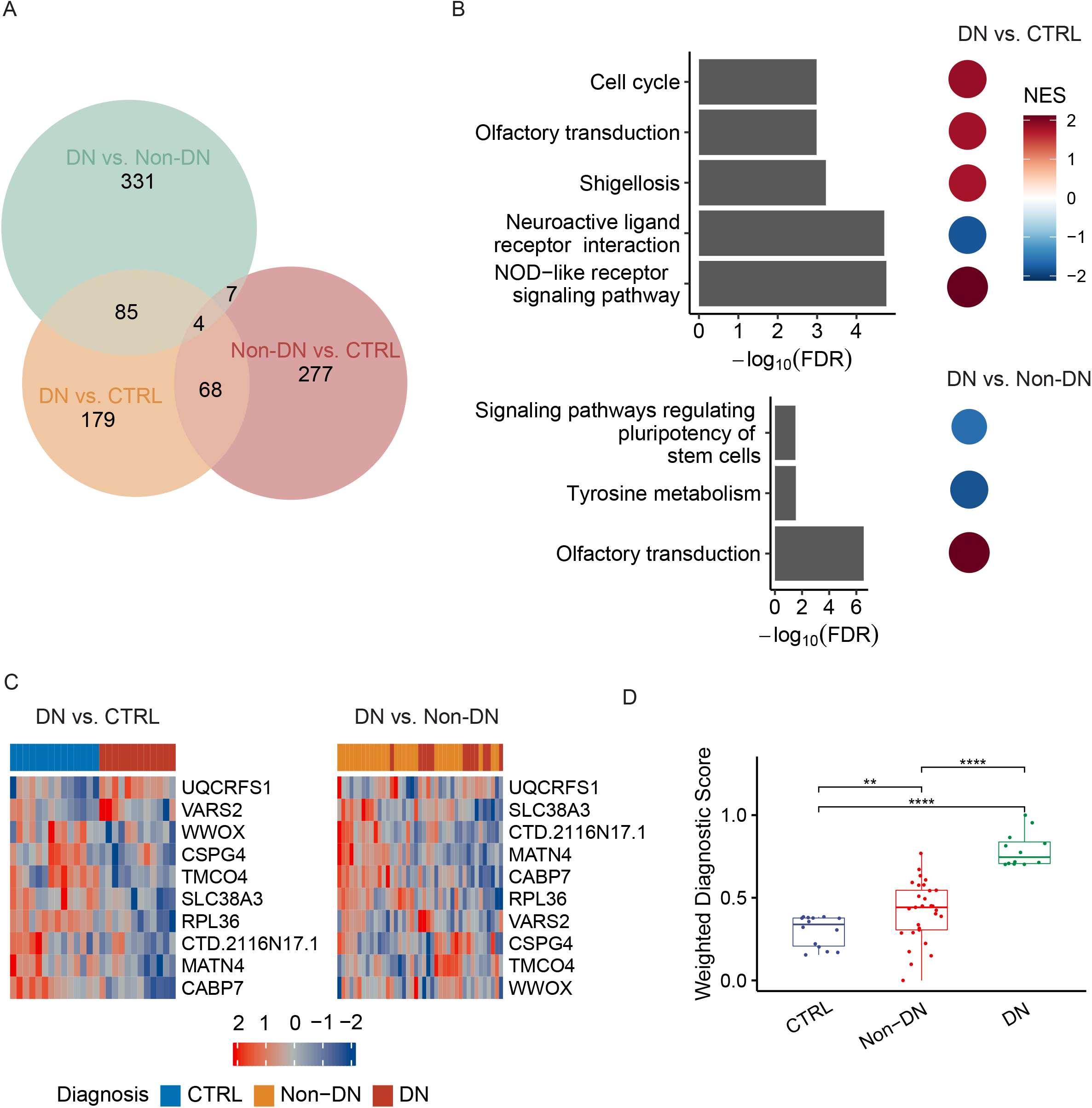
Novel 5hmC modifications implicated in diabetic nephropathy. The 5hmC-Seal data in patient-derived cfDNA reflect novel epigenetic modifications implicated in diabetic nephropathy (DN). (A). The Venn Diagram shows differentially modified gene bodies specific to DN. (B) KEGG pathways significantly enriched/depleted in DN patients relative to either controls (CTRL) or non-DN patients (Non-DN) are identified from GSEA. (C) The ten-gene panel of 5hmC marker genes shows capacity for distinguishing DN from CTRL, as well as DN from Non-DN. (D) The weighted diagnostic score (wd-score) computed with the ten-gene panel of 5hmC signature for DN shows a significant difference between DN and CRTL/Non-DN. Statistical significance: ns: p > 0.05, *: p ≤ 0.05, **: p ≤ 0.01, ***: p ≤ 0.001, ****: p ≤ 0.0001. KEGG: Kyoto Encyclopedia of Genes and Genomes; GSEA: gene set enrichment analysis; NES: normalized enrichment score.

### GSEA implicate pathways in DN-associated 5hmC

The GSEA analysis was used to identify KEGG pathways differentially modified between diagnosis groups (e.g., DN vs. controls). Specifically, the GSEA results revealed enrichment or depletion of certain canonical pathways in patients with DN relative to controls, such as NOD-like receptor signaling pathway, neuroactive ligand-receptor interaction, platelet activation, tyrosine metabolism, and necroptosis (**Figure 2B, Supplementary Table 1**). Several core genes that contributed to the enrichment or depletion of these pathways were also differentially modified between DN and T2D controls, including *CXCL1* and *PKN2* in NOD-like receptor signaling pathway, *PYY, GRM, EDN2, GCGR* and *MLN* in neuroactive ligand-receptor interaction, and *IL1B* in necroptosis (**Supplementary Table 1**). In addition, the GSEA results between DN patients and non-DN patients indicated significant enrichment or depletion of KEGG pathways such as tyrosine metabolism, olfactory transduction, and signaling pathways regulating pluripotency of stem cells (**Figure 2B, Supplementary Table 1**). However, there was no overlapping with differentially modified genes detected between DN and non-DN patients, likely due to the small sample size. Interestingly, several enriched pathways between DN and controls/non-DN patients are known to be associated with DN or kidney-related diseases, such as NOD-like receptor signaling pathway and tyrosine metabolism.(26-28)

### Summarization of a 5hmC-based wd-score for DN

A ten-gene panel of 5hmC markers (*UQCRFS1, VARS2, WWOX, CSPG4, TMCO4, SLC38A3, RPL36, CTD*.*2116N17*.*1, MATN4, CABP7*) was identified using the elastic net regularization and multivariable logistic regression models for distinguishing DN complications from T2D controls (**Figure 2C**). The wd-scores for predicting DN based on the ten-gene panel were significantly different between patients with DN complications and controls, as well as between patients with DN and those patients with non-DN complications (**Figure 2D**). When using the wd-score as the only predictor, the AUROC results showed a 100% clinical sensitivity and 97% specificity to classify DN and non-DN complication, in addition to the perfect performance of distinguishing patients with DN from controls (**Table 2**).

**Table 2.** Sensitivity and specificity of the 5hmC model and clinical variables.

| Clinical<br>Variable/Model | T2D control vs. DN |  |  | Non-DN vs. DN |  |  |
| --- | --- | --- | --- | --- | --- | --- |
|  | Sensitivity | Specificity | AUC | Sensitivity | Specificity | AUC |
| Age (year) | 0.83 | 0.50 | 0.70 | 0.42 | 0.83 | 0.54 |
| Gender<br>(male/female) | 0.75 | 0.29 | 0.52 | 0.75 | 0.55 | 0.65 |
| BMI (kg/m<sup>2</sup>) | 1.00 | 0.36 | 0.58 | 1.00 | 0.24 | 0.50 |
| Smoking (yes/no) | 0.58 | 0.57 | 0.58 | 0.58 | 0.83 | 0.71 |
| Drinking<br>(yes/no) | 0.33 | 0.86 | 0.60 | 0.33 | 0.86 | 0.60 |
| Glucose (mmol/L) | 0.73 | 0.64 | 0.68 | 0.55 | 0.68 | 0.58 |
| HbA1c (%) | 0.55 | 0.86 | 0.67 | 0.55 | 0.85 | 0.59 |
| Insulin (uU/mL) | 0.83 | 0.62 | 0.69 | 0.67 | 0.71 | 0.66 |
| CREA (umol/L) | 0.36 | 1.00 | 0.68 | 0.36 | 0.93 | 0.65 |
| UA (umol/L) | 0.64 | 0.71 | 0.60 | 0.73 | 0.48 | 0.57 |
| BUN (mmol/L) | 0.45 | 0.86 | 0.54 | 0.45 | 0.72 | 0.54 |
| eGFR<br>(mL/min/1.73m<sup>2</sup>) | 0.64 | 1.00 | 0.78 | 0.64 | 0.79 | 0.62 |
| 5hmC-based<br>Model | 1.00 | 1.00 | 1.00 | 1.00 | 0.97 | 0.98 |

Notes: BMI: Body mass index; SBP: Systolic blood pressure; DBP: Diastolic blood pressure; HbA1c: Glycated hemoglobin; CREA: Creatinine; UA: Uric acid; BUN: Blood urea nitrogen; eGFR: Estimated glomerular filtration rate. T2D: type 2 diabetes; DN: diabetic nephropathy; AUC: area under the curve.

We compared the sensitivity and specificity of various clinical variables for detecting DN from those T2D controls or patients with non-DN complications (**Table 2**). Notably, logistic regression results indicated that the 5hmC-based wd-scores in general outperformed age, gender, and various conventional clinical variables, including clinical variables or kidney functions, featuring greater AUROCs and higher sensitivity/specificity for the wd-score (**Table 2**). For example, the 5hmC-based wd-score significantly outperformed eGFR in differentiating between patients with DN and controls (AUROC: 100% vs. 78%) as well as between DN and non-DN complications (AUROC: 98% vs. 62%).

## DISCUSSION

Enhancing our understanding of molecular players of DN would provide opportunities for developing more effective clinical tools to prevent and mange this complication. In the current study, we sought to investigate DNA-associated 5hmC in patient-derived cfDNA using a cohort of T2D patients with and without DN. A substantial number of differentially modified genes were identified between T2D controls and patients with DN, as well as between patients with DN and patients with non-DN complications. Interestingly, several pathways relevant to DN such as NOD-like receptor signaling pathway, neuroactive ligand-receptor interaction, tyrosine metabolism, and necroptosis were found to be enriched in patients with DN, compared to T2D controls or patients with non-DN complications. For example, the NOD-like receptor signaling pathway and one of its member gene *CXCL1* (encoding C-X-C motif chemokine ligand 1) have been implicated in the pathogenesis and progression of DN.(26, 29) Furthermore, some significant pathways such as Fc gamma R-mediated phagocytosis and natural killer cell mediated cytotoxicity were also enriched in differential genes-associated with DN from an independent meta-analysis of mouse microarray data,(30) supporting the existence of biological links between the 5hmC landscape reflected in DN patient-derived cfDNA and the underlying pathogenesis.

Considering that cfDNA may reflect the systematic and dynamic physiological condition of a particular patient, our findings targeting novel epigenetic information in cfDNA warrant future investigations to elucidate molecular players for DN, a complex disease that is known to be affected by multiple genetic and non-genetic factors. As a matter of fact, we observed a trend of increased genome-wide 5hmC modification levels on kidney-derived enhancer marks: H3K4me1 across controls, patients with non-DN complications, and patients with DN (p-trend = 0.049). Although the current kidney-derived histone modifications included only two individuals from the Roadmap Epigenomics Project, our observation suggested that the 5hmC in cfDNA might reflect the diseased tissue in a patient with DN.

Besides differential analysis, we also sought to evaluate the possibility of whether the 5hmC profiles in cfDNA could be summarized into an integrated model to distinguish patients with DN from T2D controls as well as non-DN complications. Findings from the modeling would provide preliminary results for future development of cfDNA-based diagnostic or monitoring tools for DN. Promisingly, applying a machine-learning approach, a preliminary panel of ten genes was detected that showed high sensitivity and specificity for differentiating between patients with DN and controls/non-DN patients (**Figure 2C-D**). In particular, though limited by the sample size, the 5hmC-based model showed a trend of outperformance over various conventional clinical indexes for DN and/or diabetic complications (**Table 2**), particularly those related to kidney functions such as eGFR, strongly indicating the potential advantage of utilizing the 5hmC profiles in cfDNA as a biomarker for DN.

There are several limitations in the current study. Firstly, the sample size is relatively small. Though our primary goal was to evaluate whether novel epigenetic modifications could be detected to be implicated in DN, future larger scale investigations studies will be necessary to provide a more comprehensive picture of epigenetic landscape of DN relative to T2D controls and patients with non-DN complications. Secondly, also limited by the current sample size, our modeling of 5hmC for their diagnostic value was preliminary using a single set of samples. Though testing using patients with and without DN helped us evaluate biological relevance of the identified 5hmC features, future investigating involving more independent samples for both training and validation will be necessary to develop a clinically useful model for DN. Finally, future studies that expand to other populations and ethnic background may provide insights into any population-specific epigenetic modifications associated with DN, because of the long-appreciated baseline differences in epigenetic modifications between human populations.(31)

In conclusion, novel 5hmC distributions relevant to DN have been identified in patient-derived cfDNA samples and showed the potential of diagnostic value for DN. The 5hmC-Seal technique implemented with cfDNA holds the promise for future development of a non-invasive, clinically convenient tool for early detection of DN in high-risk T2D patients, with the ultimate goal of improving clinical outcomes through personalized preventive intervention and/or treatment.

## Data Availability

All data produced in the present study are available upon reasonable request to the authors

## AUTHOR CONTRIBUTION

Sample and data collection, clinical interpretation: YY, KY; Bioinformatics and data analysis: CZ, ZZ, QC, WZ; Funding: SL, CH, WZ; Conceptualization: CH, WZ, SL; Supervision: WZ, SL; Drafting and approval of manuscript: all authors.

## Abbreviations

BMI: Body mass index
SBP: Systolic blood pressure
DBP: Diastolic blood pressure
HbA1c: Glycated hemoglobin
CREA: Creatinine
UA: Uric acid
BUN: Blood urea nitrogen
eGFR: Estimated glomerular filtration rate
T2D: type 2 diabetes
DN: diabetic nephropathy

